# Comprehensive Quantitative Evaluation of Inter-observer Delineation Performance of MR-guided Delineation of Oropharyngeal Gross Tumor Volumes and High-risk Clinical Target Therapy: An R-IDEAL Stage 0 Prospective Study

**DOI:** 10.1101/2022.01.24.22269596

**Authors:** Carlos E. Cardenas, Sanne E. Blinde, Sweet Ping Ng, Abdallah S.R. Mohamed, Frank Pameijer, Alexis NTJ Kotte, Cornelis Raaijmakers, Brigid A. McDonald, Abrahim A. Al-Mamgani, Arash Navran, Olga Hamming-Vrieze, Nicolien Kasperts, Patricia Doornaert, Homan Dehnad, Ernst J. Smid, Andrew J. Sykes, David Thomson, Lip Wai Lee, Andrew McPartlin, Jared Robbins, Kate Newbold, Chris Nutting, Irene Karam, Ian Poon, Marielle Philippens, Hesham Elhalawani, Mona J Kamal, Mohamed AM Meheissen, Adam Garden, William H. Morrison, Jack Phan, G. Brandon Gunn, Steven J. Frank, David I Rosenthal, Pierre Blanchard, Houda Bahig, Clifton D. Fuller, Chris Terhaard

## Abstract

**Purpose:** Tumor and target volume manual delineation remains a challenging task in head and neck cancer radiotherapy. The purpose of this study is to conduct a multi-institutional evaluation of manual delineations of gross tumor volume (GTV), high-risk clinical target volume (CTV), parotids, and submandibular glands on treatment simulation MR scans of oropharyngeal cancer (OPC) patients.

**Methods:** Pre-treatment T1-weighted (T1w), T1-weighted with Gadolinium contrast (T1w+C) and T2-weigted (T2w) MRI scans were retrospectively collected for 4 OPC patients under an IRB-approved protocol. The scans were provided to twenty-six radiation oncologists from seven international cancer centers who participated in this delineation study. In addition, each patient’s clinical history and physical examination findings along with a medical photographic image and radiological results were provided. The contours were compared using overlap and distance metrics using both STAPLE and pair-wise comparisons. Lastly, participants completed a brief questionnaire to assess personal experience and CTV delineation institutional practices.

**Results:** Large variability was measured between observers’ delineations for both GTVs and CTVs. The mean Dice Similarity Coefficient values across all patients’ delineations for GTVp, GTVn, CTVp, and CTVn where 0.77, 0.67, 0.77, and 0.69, respectively, for STAPLE comparison and 0.67, 0.60, 0.67, and 0.58, respectively, for pair-wise analysis. Normal tissue contours were defined more consistently when considering overlap and distance metrics. The median radiation oncology clinical experience was 7 years and the median experience delineating on MRI was 3.5 years. The GTV-to-CTV margin used was 10 mm for six of seven participant institutions. One institution used 8 mm and three delineators (from three different institutions) used a margin of 5 mm.

**Conclusion:** The data from this study suggests that appropriate guidelines, contouring quality assurance sessions, and training are still needed for the adoption of MR-based treatment planning for head and neck cancers. Such efforts should play a critical role in reducing inter-observer delineation variation and ensure standardization of target design across clinical practices.

**Summary:** With the rapid increase of MR imaging use in radiotherapy, MRI presents with endogenous contrast greater than CT for tumor and target definition; however, reports on delineation variability for these volumes on MRI are limited. In this prospective manual segmentation challenge of 26 radiation oncologists, we formally quantify human performance and heterogeneity in physician tumor and target delineations based on MRI scans of oropharyngeal cancer patients.

## Introduction

The widespread adoption of highly conformal therapies such as intensity modulated radiation therapy (IMRT) and stereotactic body radiation therapy (SBRT) for head and neck cancer treatment has resulted in improved sparing of organs at risk and has reduced the toxicity burdens typically associated with radiation therapy. While the clinical benefits of these techniques are well documented [3, 12, 21], the use of highly conformal plans brought about new challenges to the clinic [7]. With the use of high precision treatments there has been a larger focus on accurate target delineation, patient set-up, and treatment delivery since small errors while performing these tasks may results in significant under-dosage of at-risk regions and/or over-dosage of surrounding organs at risk (OARs).

The delineation of tumor and target volumes have been greatly improved by the adaptation of multi-modality imaging in radiation oncology. It is common clinical practice to use a contrast-enhanced computed tomography (CE-CT) with or without a fluorodeoxyglucose positron emission tomography (FDG-PET) scan for head and neck cancers as they greatly improve the ability to see macroscopic tumor involvement over non-contrast CT alone. More recently, magnetic resonance imaging (MRI) has become more widely used in radiotherapy due to their higher soft tissue contrast over CT, which often allows for better distinction between healthy tissues and appreciable disease. Furthermore, with the advent of the MR-Linac[1]–[3] and MR-guided radiation therapy (MRgRT), there is a trend toward a MR-based radiation treatment planning[4], [5] increasing the likelihood of future MR-based tumor and target volume delineation.

Inter- and intra-observer variability when delineating gross tumor volumes (GTV) and clinical target volumes (CTV) have been widely studied for many treatment sites with many reports suggesting large heterogeneity amongst practitioners. This large variability in target delineation is considered a major source of uncertainty [6], [7] and reduces our ability to systematically assess the quality of the radiation therapy plans. The inter-observer variability for delineation of the CTV for oropharyngeal cancer is one of the largest reported in the literature [7]. When delineating tumors alone, Thiagarajan et al [8] investigate the contributions of MRI and FDG-PET on forty-one head and neck cancer patients and found improved agreement when using multi-modality information over single modality alone; in addition, they found that the lack of physical examination (PE) findings resulted in an underestimation of mucosal disease when cases were presented without knowledge of PE findings. Focusing on oropharyngeal cancers, Bird et al [9] found that inter-observer delineation variability was higher when using CT-alone than both MR-alone and CT+MR. Similar results have been reported by Rasch et al [10] for nasopharynx tumors. Hong et al [11] conducted a study to assess this variability on an oropharyngeal cancer patient and noticed significant variability in target delineation and clinical practices. While several head and neck target delineation guidelines have been published in recent years [12]–[17] these guidelines focus on CT-based radiotherapy and may not be suitable for MR-based radiation treatment planning. Furthermore, MR-based CTV inter-observer delineation variability is currently unknown for oropharyngeal cancers.

The MR-LinAc Consortium is a multi-site cooperative group[18], committed to prospective technology development in a programmatic format, using a paradigm based on the surgical technology IDEAL (Idea, Development, Exploration, Assessment, Long-term study) conceptual framework, deemed R-IDEAL [19] (Radiotherapy-Idea, Development, Exploration, Assessment, Long-term study). As part of this effort in preparation for now-open Phase II adaptive MR-guided radiotherapy trial for oropharyngeal cancer[20], the MR-Linac Consortium Head and Neck Tumor Site Group sought to undertake a prospective technical benchmarking evaluation (R-IDEAL Stage 0) of human segmentation performance, as part of a coherent quality assurance program[21] for multisite MR-LinAc trials[20].

Consequently, the aim of this prospective, blinded R-IDEAL Stage 0 technology implementation study was to 1) quantify observer-dependent inter-observer manual segmentation variability for GTVs and high-risk CTVs as a necessary reprequisite for adaptive trial that modify tumor volumes on the MR-Linac, as well as index organs-at-rsik (parotid and submandibular glands (SMG)), for oropharyngeal cancer patients using magnetic resonance imaging inputs.

## Methods and Materials

### Patients and Image Acquisition

Four patients with oropharyngeal cancer were retrospectively selected by two experienced head and neck radiation oncologists from different centers after receiving institutional review board approval. Patients with both early and locally advanced stage disease were selected. Case characteristics are shown in Table 1.

**Table 1.** Case characteristics

|  | CASE 1 | CASE 2 | CASE 3 | CASE 4 |
| --- | --- | --- | --- | --- |
| **PRIMARY SITE** | Left tonsil | Base of tongue right-sided | Posterior wall | Base of tongue bilateral |
| **T-STAGE** | 1 | 2 | 3 | 3 |
| **N-STAGE** | 0 | 0 | 2b | 1 |
| **P16 STATUS** | + | + | - | + |
| **SMOKING STATUS** | Non-smoker | Non-smoker | Smoker | Smoker |

Pre-treatment T1-weighted (T1w), T1-weighted with Gadolinium contrast (T1w+C) and T2-weigted (T2w) MRI scans were available for all patients. These scans were acquired on an Ingenia 3T MRI scanner (Philips, Eindhoven, The Netherlands) for treatment planning purposes with each patient in the treatment planning position and using a thermoplastic mask and an immobilization device. The scans covered the region extending from the caudal-edge of the nasopharynx region cranially and the hypopharynx region caudally in the superior– inferior direction, respectively. Details on image acquisition are presented in Table 2.

**Table 2.** Summary of magnetic resonance image acquisition parameters

|  |  | Case 1 | Case 2 | Case 3 | Case 4 |
| --- | --- | --- | --- | --- | --- |
| **T1w and T1w+C** | **Acquisition Sequence** | 2D TSE mDixon | 2D TSE mDixon | 2D TSE mDixon | 2D TSE CLEAR |
|  | **Flip Angle** | 90° | 90° | 90° | 90° |
|  | **Repetition Time** | 742 ms | 742 ms | 742 ms | 597 ms |
|  | **Echo Time** | 14 ms | 14 ms | 14 ms | 10 ms |
|  | **Echo Train Length** | 8 | 8 | 8 | 10 |
|  | **N Averages** | 2 | 2 | 2 | 2 |
|  | **Field of View** | 280 x 280 mm<sup>2</sup> | 200 x 200 mm<sup>2</sup> | 280 x 280 mm<sup>2</sup> | 200 x 200 mm<sup>2</sup> |
|  | **Matrix Size** | 384 x 384 | 256 x 256 | 384 x 384 | 400 x 400 |
|  | **Pixel Spacing** | 0.729 mm | 0.782 mm | 0.729 mm | 0.5 mm |
|  | **Slice Spacing** | 2 mm | 2 mm | 2 mm | 1 mm |
| **T2w** | **Acquisition Sequence** | 2D TSE mDixon | 2D TSE mDixon | 2D TSE mDixon | 3D VISTA SENSE |
|  | **Flip Angle** | 90° | 90° | 90° | 90° |
|  | **Repetition Time** | 4176 ms | 4176 ms | 4176 ms | 1400 ms |
|  | **Echo Time** | 100 ms | 100 ms | 100 ms | 206 ms |
|  | **Echo Train Length** | 22 | 22 | 22 | 85 |
|  | **N Averages** | 2 | 2 | 2 | 2 |
|  | **Field of View** | 280 x 280 mm<sup>2</sup> | 200 x 200 mm<sup>2</sup> | 280 x 280 mm<sup>2</sup> | 250 x 250 mm<sup>2</sup> |
|  | **Matrix Size** | 576 x 576 | 400 x 400 | 576 x 576 | 512 x 512 |
|  | **Pixel Spacing** | 0.486 mm | 0.5 mm | 0.486 mm | 0.488 mm |
|  | **Slice Spacing** | 2 mm | 2 mm | 2 mm | 0.75 mm |

TSE: Turbo Spin Echo, mDixon: multiple-point Dixon, CLEAR: Constant LEvel AppeaRance, VISTA: Volume ISotropic Turbo spin echo Acquisition, SENSE: SENSitivity Encoding

**Table 3.**
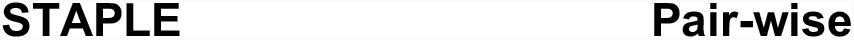
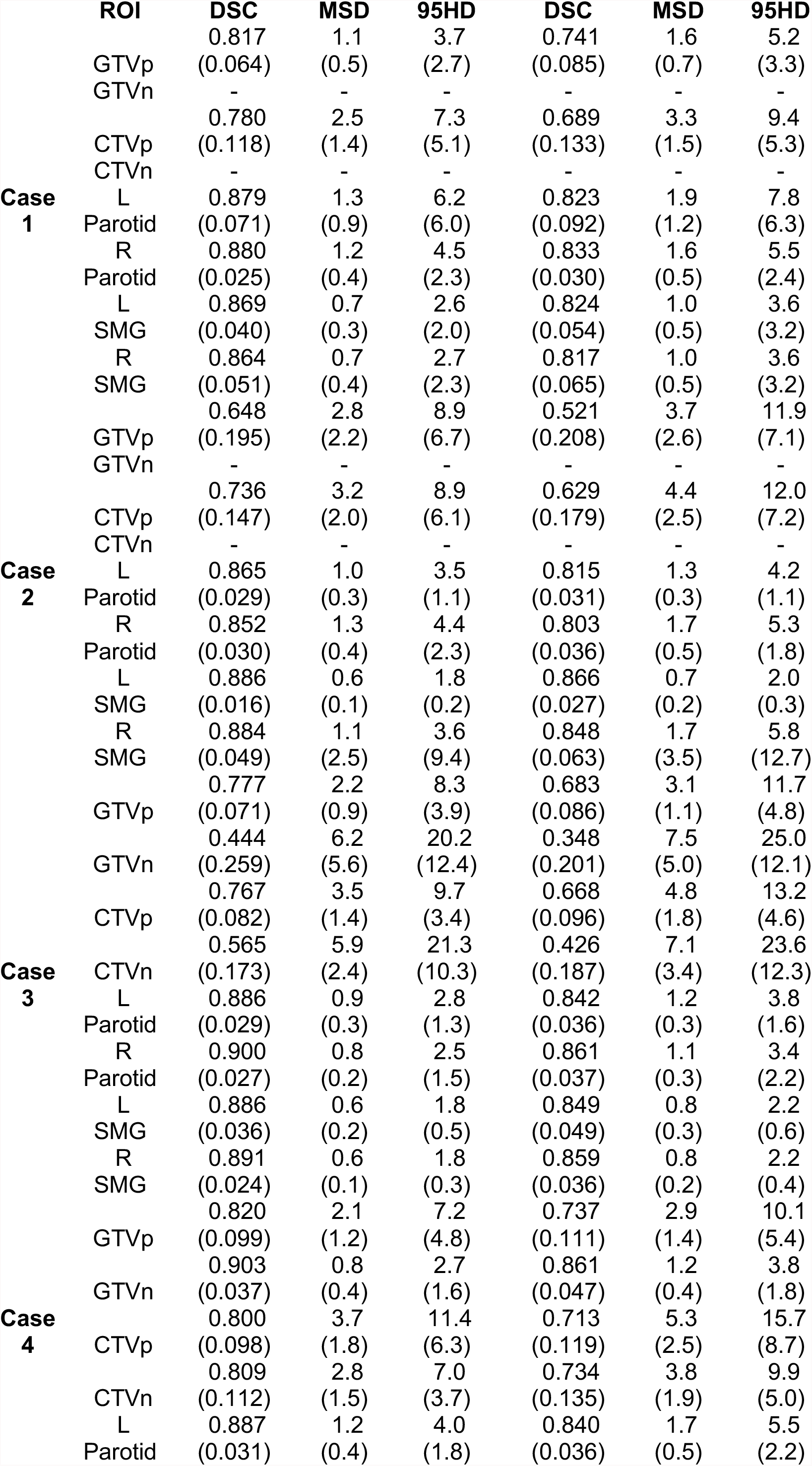
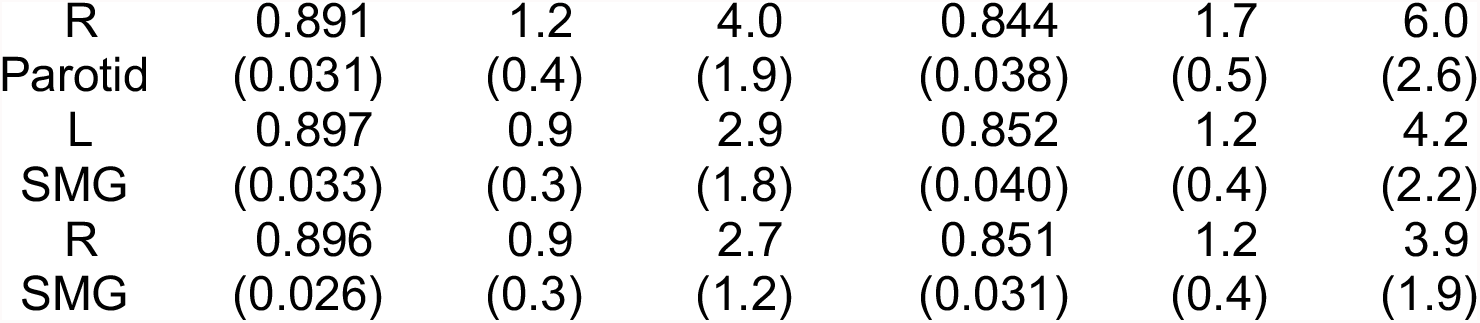
Summary of calculated average (standard deviation) values for the Dice Similarity Coefficient (DSC), mean surface distance (MSD), and 95^th^ percentile Hausdorff distance (95HD) for both STAPLE and Pair-wise comparisons.

### Delineation Study

Twenty-six radiation oncologists and one dedicated head and neck radiologist from seven international centers (UMC Utrecht (The Netherlands), University of Texas MD Anderson Cancer Center (Houston, Texas, USA), NKI Antoni van Leeuwenhoek (Amsterdam, The Netherlands), Sunnybrook Health Sciences Centre (Toronto, Ontario, Canada), Froedtert & Medical College of Wisconsin Cancer Center (Milwaukee, Wisconsin, USA), The Royal Marsden NHS Foundation Trust (London, UK), The Christie NHS Foundation Trust, (Manchester, UK)) were asked to delineate the parotids, submandibular glands, the GTV and high-risk CTV. When nodal disease was present, participants were asked to delineate GTVp/CTVp and GTVn/CTVn as separate structures to investigate delineation differences between primary and nodal disease regions.

The available pre-treatment MRI scans (T1w, T1w+C, and T2w) were provided with each patient’s clinical history and physical examination findings along with a medical photographic image and radiological results. Participants were asked to delineate the requested structures based on their own institutional guidelines. In addition, all participants received a basic questionnaire to determine years of experience in radiotherapy, years of experience with delineating on MRI, delineation software, software settings and institutional GTV-to-CTV margin expansion values used.

### Quantitative Analysis

The contours delineated in this study were compared to quantify inter-observer variability. The Dice similarity coefficient[22] (DSC), the mean surface distance (MSD), and the 95th Percentile Hausdorff distance (95HD) where calculated. These metrics are defined as follows,

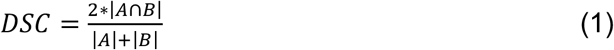

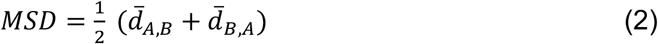

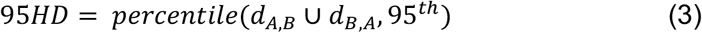

where |A| and |B| are the number of voxels from contoured volumes A and B, respectively; |A∩B| denotes the number of voxels included in the intersection between volumes A and B; *d*_*A,B*_ is a vector containing all minimum Euclidian surface distances from the surface point from volume A to B; and 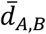 is the average value in the vector *d*. The DSC ranges in values from 0 (no overlap) to 1 (perfect overlap); for both MSD and 95HD, values closer to zero represent better agreement between two contours’ surfaces.

These metrics (Eqs. 1-3) were calculated to assess the manual delineations using two approaches (Figure 1). First, a physician pair-wise comparison of the contours was performed, meaning that all physician comparisons (i.e. Physician 1 vs Physician 2, Physician 1 vs Physician 3, …, Physician 25 vs Physician 26) were considered. This comparison provides a real-world estimate of the delineation variability amongst the participants providing the minimum and maximum extreme derived from the overlap and distance metrics. Second, we estimated a consensus volume using a modified version [23] of the simultaneous truth and performance level estimation (STAPLE) algorithm [24]. The STAPLE algorithm calculates the maximum likelihood estimates of the true positive and false negative of individual segmentations and uses these values to produce a volume that estimates the best agreement between the individual segmentations. A limitation to the STAPLE algorithm is that it does not take into consideration intensity information of the image to be segmented, it only relies on individual segmentations. The methodology by Yang et al [23] addresses this limitation by creating a tissue appearance model and integrating it into the STAPLE fusion process. The resulting consensus volumes for each patient were considered our ground-truth for this analysis and we compared each physician’s delineations using overlap (DSC) and distance (MSD and 95HD) metrics.

**Figure 1.**
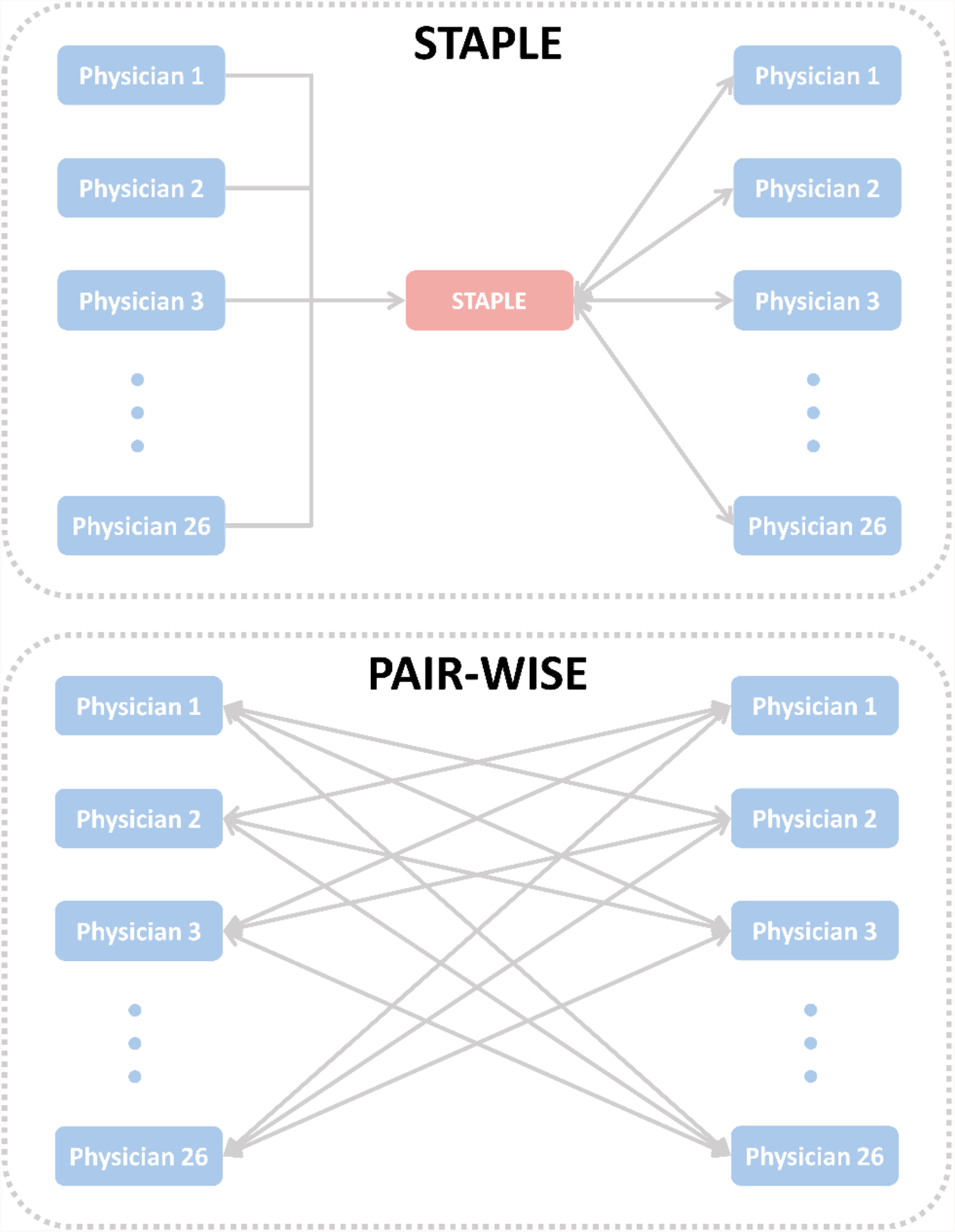
Methods for quantitative evaluation of manual contours. The STAPLE algorithm was used to generate a “consensus” contour (top) for each organ, tumor, and target volume contours; then individual physician contours are compared to the individual region of interest’s STAPLE contour. In addition, a pair-wise evaluation of the contours was performed (bottom); here, individual physician contours are individually compared to every other physician contour. This approach highlights the potential true disagreement between two individual physicians.

## Results

Twenty-four out of twenty-six (92%) radiation oncologists submitted a full set of contours for all patients. Incomplete submissions from 2 participants led to the exclusion of all contours from these participants in the subsequent analysis. The head and neck radiologist successfully submitted GTVp and GTVn contours for all patients; these contours were taken into consideration in the analysis, but were not used to generate any STAPLE volumes. Twenty-two of twenty-six radiation oncologists’ questionnaires were completed. The median time of experience as a head and neck radiation oncologist was 7 years (range: 1-25 years). The median time of experience with delineating on MRI was 3.5 years (range: 0-15 years). Three observers used automatic segmentation with manual edits to delineate the organs at risk. The GTV-to-CTV margin used was 10 mm for six of seven participant institutions. One institution used 8 mm and three delineators (from three different institutions) used a margin of 5 mm.

Volumes (in cm^3^) for the participant’s delineations and resulting STAPLE volumes are detailed in Supplementary Tables S1 and S2. The average volumes (± standard deviation) for GTVp for cases 1 through 4 were 6.5 cm^3^ (± 1.4), 8.0 cm^3^ (± 5.4), 32.8 cm^3^ (± 12.2), and 21.0 cm^3^ (± 6.7), respectively; GTVn mean volumes for cases 3 and 4 were 5.2 cm^3^ (± 5.3) and 7.9 cm^3^ (± 1.0). The STAPLE volumes for GTVp and GTVn were 6.1 cm^3^, 7.6 cm^3^, 32.5 cm^3^, and 21.8 cm^3^, for cases 1 through 4, respectively, and 2.1 cm^3^ and 7.6 cm^3^ for cases 3 and 4. In regards to CTVp and CTVn, average volumes were 36.4 cm^3^ (± 13.7), 40.2 cm^3^ (± 21.1), 117.7 cm^3^ (± 51.9), and 79.5 cm^3^ (± 29.9), for cases 1 through 4, respectively, and 33.7 cm^3^ (± 23.4) and 36.2 cm^3^ (± 11.0) for cases 3 and 4. Considering normal tissues, the average left and right parotid volumes were 26.4 cm^3^ (± 4.0), 17. cm^3^ (± 2.1), 23.1 cm^3^ (± 2.6), 27.7 cm^3^ (± 3.5) and 24.8 cm^3^ (± 3.2), 16.9 cm^3^ (± 2.4), 22.8 cm^3^ (± 1.9), 27.4 cm^3^ (± 3.0), respectively, for cases 1 through 4; their corresponding STAPLE volumes were 25.6 cm^3^, 16.3 cm^3^, 22.8 cm^3^, 28.2 cm^3^ and 23.3 cm^3^, 15.5 cm^3^, 22.1 cm^3^, and 28.2 cm^3^, respectively. The average left and right submandibular volumes were 6.7 cm^3^ (± 1.0), 7.0 cm^3^ (± 0.5), 7.6 cm^3^ (± 0.6), 12.6 cm^3^ (± 1.3) and 6.5 cm^3^ (± 1.0), 6.6 cm^3^ (± 1.0), 8.0 cm^3^ (± 0.5), 12.8 cm^3^ (± 1.1), respectively, for cases 1 through 4; their corresponding STAPLE volumes were 6.5 cm^3^, 6.2 cm^3^, 7.1 cm^3^, 12.7 cm^3^ and 6.3 cm^3^, 6.1 cm^3^, 7.6 cm^3^, and 12.7 cm^3^, respectively. The median coefficient of variation (standard deviation / mean × 100%) across all cases for CTVs, GTVs, parotids, and submandibular glands were 40.9% (range: 30.4 – 69.5%), 34.5% (range: 12.2 – 101.0%), 12.5% (range: 8.5 – 14.9%), and 9.8% (range: 6.4 – 15.4%), respectively.

Boxplots displaying distributions from the volumetric comparisons using STAPLE and pair-wise evaluation approaches are shown in Figures 1 and 2 for GTVs/CTVs and normal tissues, respectively. A summary of these results is provided in Table 2. When considering primary and nodal volumes, both GTVp and CTVp delineations were found to be more variable than GTVn and CTVn delineations (p-values: 0. 01 and < 0.0001, respectively) when comparing their respective DSC values (measured against STAPLE). When considering laterality of the normal tissues, there was no significant difference in DSC distributions from left and right manual delineations (measured against STAPLE).

**Figure 2.**
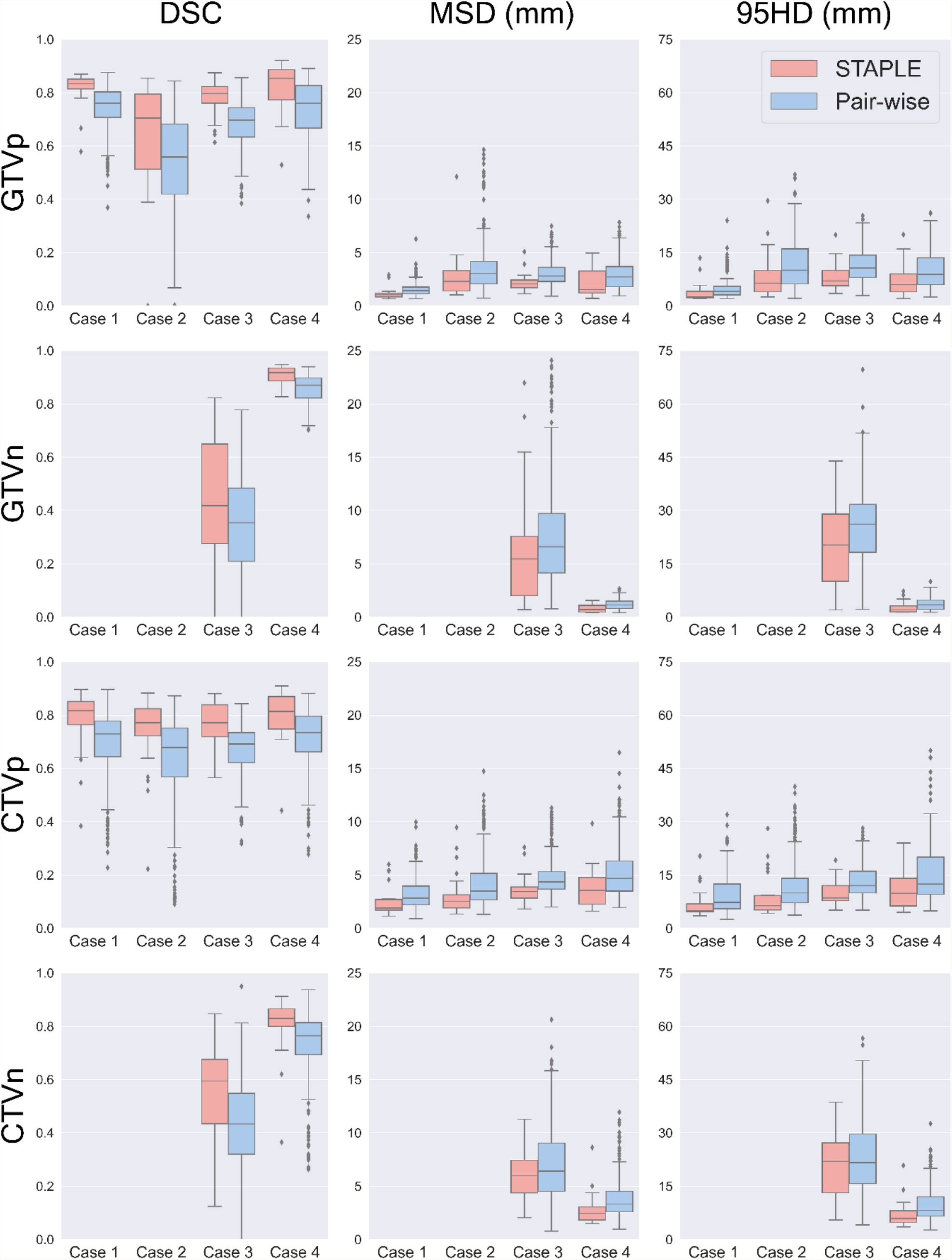
Boxplots demonstrating inter-observer variability for GTV and CTVs, shown in rows, for the Dice Similarity Coefficient (DSC), mean surface distance (MSD), and 95^th^ percentile Hausdorff distance (95HD), shown in columns. All distances are in millimeters.

## Discussion

This study presents the results of a numerically robust R-IDEAL Stage 0 multi-institutional quantitifcation of inter-observer segmentation/delineation variability of gross tumor volumes, high-risk clinical target volumes, parotids, and submandibular glands of oropharyngeal cancer patients when these structures are delineated on MR images alone (as would be the vcase for daily MR-LinAc-based adaptive MR-guided-radiotherapy. The data suggests substantial variability in the delineation of gross tumor and clinical target volumes across participants. For example, ratio in volumes between smallest and largest volumes (V_max_/V_min_) across all participants were as high as 31.0 for tumor volumes and 32.3 for target volumes (average across all cases: 10.7 and 11.4 for GTV and CTV, respectively; see Supplementary Data). Figure 4 shows individual participant’s delineations (center columns) for GTVs and CTVs on single axial T1w MR scan slices for the 4 cases presented in this study. In this figure, the right most panels (“CTV*”) show axial and sagittal or coronal views of the STAPLE contours for GTV and CTV, as well as the intersection and union of all participant’s CTVs. Interestingly, for 4 out 6 GTVs, the consensus GTV contours (STAPLE) were mostly covered by the intersection of all participant’s target volumes suggesting that the consensus derived GTV would receive appropriate coverage by all participant’s CTVs; however, for 2 cases the CTV intersection volume had little overlap with the STAPLE GTVs with one of these cases showing that there was not a single voxel in the patient’s MR scan where 100% of participants CTVs overlapped (Figure 4, panels denoted by asterisk).

There was higher agreement in the delineation of normal tissues with ratios in volumes across participants being as high as 2.8 for parotids and 2.2 for submandibular glands (average across all cases: 1.9 and 1.6 for parotids and submandibular glands, respectively). Figure 3 shows the STAPLE and pair-wise comparison of inter-observer delineations per case. It is important to note that one participant contoured the left submandibular gland as the right submandibular gland resulting in zero overlap (this contour was excluded from analysis and Figure 3) between this organ’s contour and the remaining delineations, highlighting the need for quality assurance of the contours.

**Figure 3.**
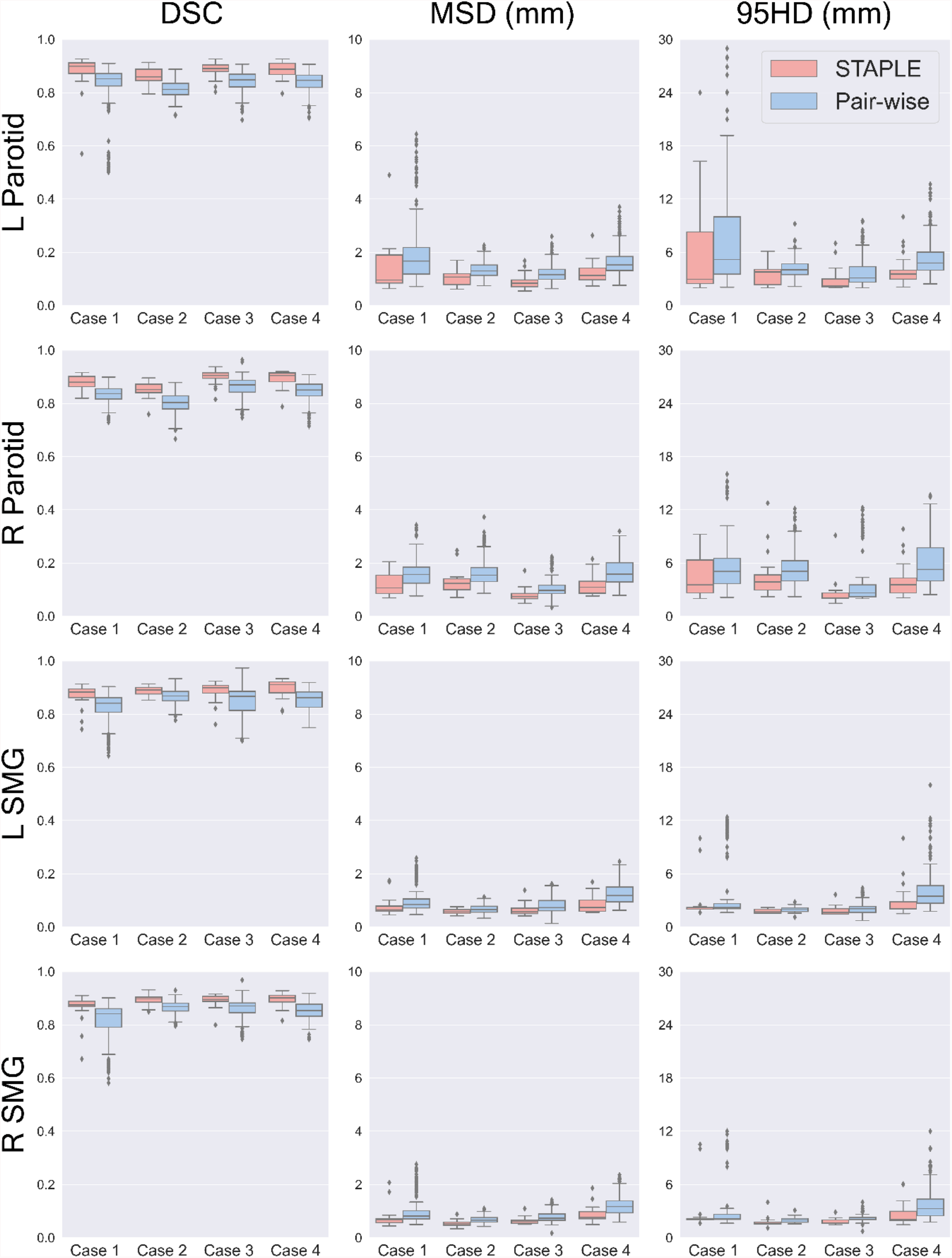
Boxplots demonstrating inter-observer variability for parotids and submandibular glands (SGMs), shown in rows, for the Dice Similarity Coefficient (DSC), mean surface distance (MSD), and 95^th^ percentile Hausdorff distance (95HD), shown in columns. All distances are in millimeters.

**Figure 4.**
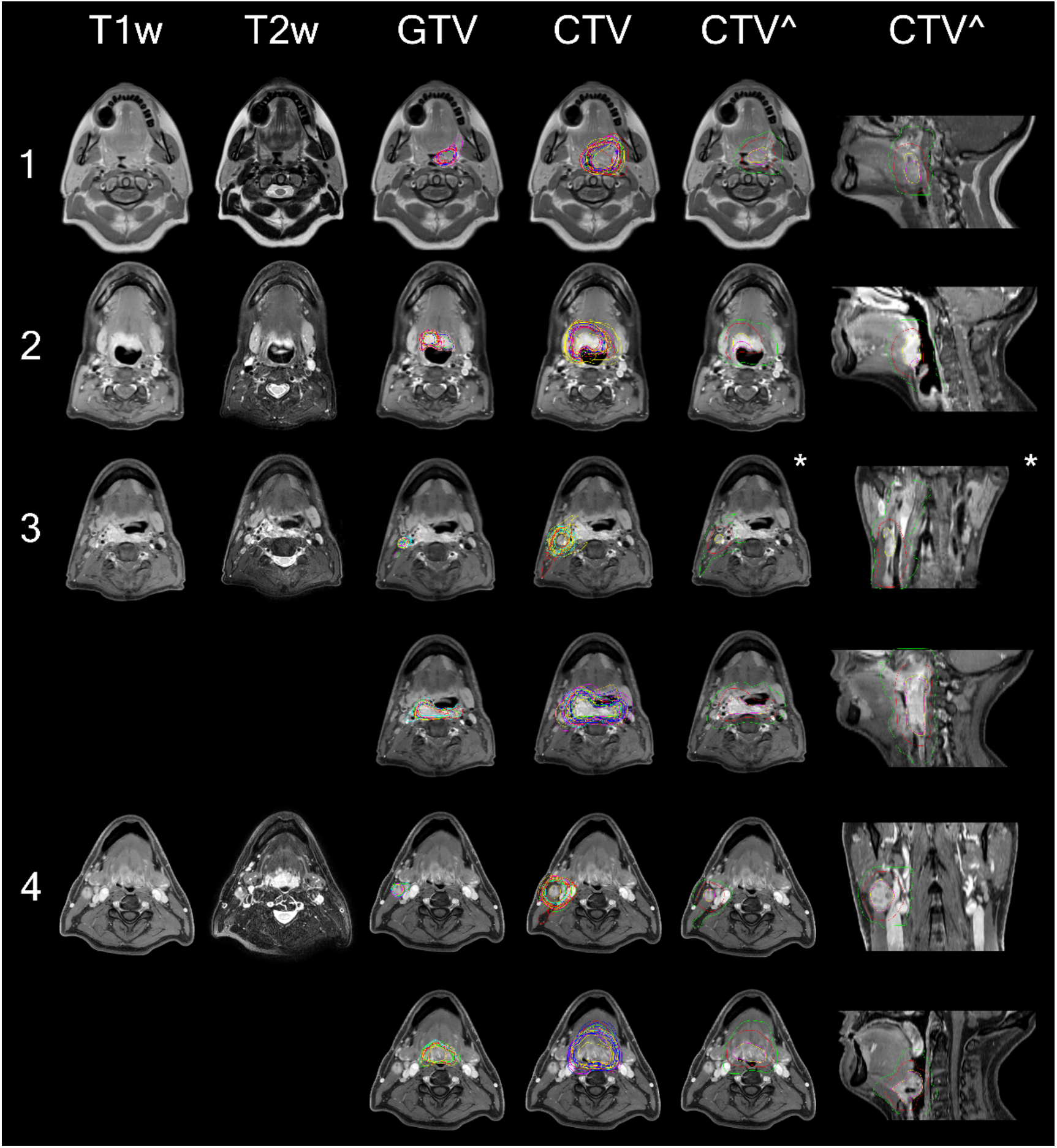
Illustration demonstrating delineation variability for both GTV and CTV in all 4 cases (rows). For cases 3 and 4, nodal disease is shown on their respective first rows then followed by primary disease and target delineations on the following row panels. From left to right each column shows a) an axial slice of the T1w+C MRI (T1w+C) scan provided, b) an axial slice of the T2w MRI (T2w) scan provided, c) all participants’ GTV contours (GTV), d) all participants’ CTV delineations (CTV), e) axial view of STAPLE contours for GTV (yellow) and CTV (red), union of all CTV contours (green), and the intersection of all CTV contours (fuchsia), f) sagittal/coronal view from e).

Two approaches were used to quantitatively evaluate agreement between participant’s contours: 1) individual contour comparison to consensus (generated via STAPLE) and 2) pair-wise comparison of the contours. The STAPLE algorithm generates a statistically-derived “consensus” volume by taking into consideration delineations from multiple observers. While the generation of a computationally-derived “consensus” can be attractive for inter-observer analysis, real-clinical scenarios lack “consensus” volumes; therefore, it could be argued that pair-wise analyses may provide a more accurate estimate of inter-observer variability. Our data showed that the STAPLE comparison distributions where tighter than those observed for the pair-wise comparison distributions for both tumor/target volumes and normal tissues (Figures 2 and 3, respectively). This was expected as the pair-wise comparison provides a more accurate quantification of extreme differences in contours between two participants (i.e. these differences are lessened by comparing these extremes to a common contour in the STAPLE analysis).

Several studies have reported large inter-observer delineation variability for head and neck GTV and CTVs [8], [11], [25]–[30]. Anderson et al [26] investigated head and neck GTV delineation variability across multiple imaging modalities (CE-CT, FDG-PET/CT, and T1w+C MRI). MRI-based GTVs resulted, on average, in the largest delineated volumes, with an average intersection over union of 36% across three observers. In a similar study, Ng et al [28] found delineations based on T1w+C MR imaging alone resulted in less GTV inter-observer delineation variability (median DSC of 0.58 in a pair-wise analysis) when compared to dual-energy CT. Gudi et al [29] reported moderate GTV delineation variability on both CE-CT and CE-CT + FDG-PET/CT (mean DSC values of 0.57 and 0.69, respectively) measured on 10 cases with pharyngolaryngeal cancer. Similarly, Thiagarajan et al [8] showed significant variability in GTV delineation when evaluating the individual contributions of MRI, PET, and physical evaluation in the delineation process. Concerning CTVs, Hong et al previously showed large heterogeneity in target design between different observers [11]. In their study, the authors provided participants (n=20) with CT scans and GTV contours of an oropharyngeal cancer patient (T2 N1 M0 squamous cell carcinoma of the tonsil) and asked the participants to delineate CTVs. When considering high-risk CTVs, the coefficient of variation in their study was 191% (w/ μ = 43 cm^3^ and σ = 82 cm^3^) showing larger variation than the current study (CoV = 41%). To address this reported variability in CTV delineation, some groups have proposed the use of uniform margin expansions [31] or computational methods for the automatic delineation of CTVs [32]– [34]. Hansen et al [31] showed in a multi-center study that using geometric margins from GTV-to-CTV for high-risk CTVs resulted in higher agreement in manual delineations than when using anatomical margins. Cardenas et al [32] proposed the use of artificial intelligence to automatically delineate high-risk CTVs. Their results showed high agreement between the clinically-used and automatically-delineated target volumes (mean DSC = 0.81 vs mean DSC = 0.64 in the current inter-observer pair-wise analysis) when GTVs are already provided. In the current study, we noticed slight improvement in terms of consistency for primary GTV and CTV contours when compared to those from base of tongue cases, this may be caused by the fact that lymphoid tissue is abundantly present in this region and might be misinterpreted as tumor on MRI, but additional analyses are needed to confirm this hypothesis. Consequently, future oropharyngeal cancer delineation studies should consider techniques to integrate microscopic disease evaluation to provide margin guidelines to the community. For example, Ligtenberg et al [35] proposed imaging modality-specific margins for laryngeal and hypopharyngeal cancer after co-registering pre-surgical CT, MRI, and FDG-PET imaging with histological images collected after laryngectomy. Nevertheless, similar studies for oropharyngeal cancers would bring additional challenges.

The use of computed tomography scans for radiotherapy treatment planning has been required as these scans provide electron density information that is necessary for previously clinically-available dose calculation algorithms. With the introduction of MRgRT and advances in MR-based dose calculation algorithm development, our field is fast approaching the possibility of MR-based radiotherapy. There are many advantages to MR-based radiotherapy. For example, the assessment of head and neck cancers, particularly those located in the oropharynx, can be hindered due to several factors including the presence of CT dental artifacts and lack of contrast between tumor and surrounding tissues. Several studies have shown MRI to provide superior soft tissue contrast allowing for better definition of tumor extent and adjacent organs at risk [36], [37]. Furthermore, the use of single modality scans for treatment planning removes the need to co-register images eliminating any potential treatment uncertainties derived from image co-registration. It goes without saying that the introduction of MR-based radiotherapy presents itself with unique challenges. The results of the current study suggest that there is an urgent need for MR-based delineation guidelines and training necessary to reduce inter-observer delineation variability of tumor and target volumes. It is important to highlight the potential role of conducting contouring peer-review sessions prior to treatment commencement during initial adoption of MRgRT in a clinical setting. Some studies have suggested that establishing institutional contouring peer-review sessions leads to more consistent target design within institutional clinical practices [38], [39].

The presented study is subject to some limitations. First, only images from T1w, T1w+C, and T2w scans were provided to participants for contouring. Some studies have indicated that the addition of fluorodeoxyglucose positron emission tomography (FDG-PET/CT) and/or contrast-enhanced CT (CE-CT) to MR scans could produce more consistent tumor delineations. Furthermore, it does not consider the role of multi-parametric MR imaging as a venue to provide additional information about tumor disease and extent. Future studies will be needed to determine the role and potential benefit of including additional imaging modalities for delineation purposes. A follow-up study is underway to investigate the addition of FDG-PET imaging and the use of recommended guidelines for GTV definition using MRI [40] which may lead to better delineation conformity between observers.

Importantly, this study represents, to our knowledge, not only the 1^st^ formal prospective technical assessment of MR-only radiotherapy segmentation required to benchmark performance for MR-guided adaptive radiotherapy, but also the largest single multi-site target delineation cohort for oropharyngeal cancers. This robust sample size provides a statistically reliable estimator of segmentation agreement, using standard measurands of interobserver performance, we found that MR-only performance appears to meet or exceed agreement and consistency metrics for prior head and neck target delineation series for GTV, CTV, and OARs tested. Consequently, we feel confident that this effort provides a formal justification and quantitative benchmark, and provides a compelling rationale for adaptive GTV/CTV modification on clinical trials, or GTV/CTV/OAR monitoring as standard of care on MR-LinAc devices enabled with sequences comparable to those listed.

## Conclusion

Tumor and target volume manual delineation remains a challenging task in head and neck cancer radiotherapy. The data from this study suggests that appropriate guidelines, contouring quality assurance sessions, and training are still needed for the adoption of MR-based treatment planning for head and neck cancers. Such efforts should play a critical role in reducing inter-observer delineation variation and ensure standardization of target design across clinical practices.

## Data Availability

Anonymized data are embargoed until peer-review completion, but thereafter available at https://doi.org/10.6084/m9.figshare.18822194

## References

[1] J. J. W. Lagendijk, B. W. Raaymakers, A. J. E. Raaijmakers, et al., “MRI/linac integration,” Radiother. Oncol., vol. 86, no. 1, pp. 25–29, Jan. 2008.

[2] B. W. Raaymakers, J. J. W. Lagendijk, J. Overweg, et al., “Integrating a 1.5 T MRI scanner with a 6 MV accelerator: proof of concept,” Phys. Med. Biol., vol. 54, no. 12, pp. N229–N237, 2009.

[3] S. Mutic and J. F. Dempsey, “The ViewRay System: Magnetic Resonance–Guided and Controlled Radiotherapy,” Semin. Radiat. Oncol., vol. 24, no. 3, pp. 196–199, Jul. 2014.

[4] M. A. Schmidt and G. S. Payne, “Radiotherapy planning using MRI,” Phys. Med. Biol., vol. 60, no. 22, pp. R323–R361, Nov. 2015.

[5] C. Kontaxis, G. H. Bol, B. Stemkens, et al., “Towards fast online intrafraction replanning for free-breathing stereotactic body radiation therapy with the MR-linac,” Phys. Med. Biol., vol. 62, no. 18, pp. 7233–7248, Aug. 2017.

[6] M. Van Herk, “Errors and Margins in Radiotherapy,” Semin. Radiat. Oncol., vol. 14, no. 1, pp. 52–64, 2004.

[7] B. Segedin and P. Petric, “Uncertainties in target volume delineation in radiotherapy -Are they relevant and what can we do about them?,” Radiol. Oncol., vol. 50, no. 3, pp. 254–262, 2016.

[8] A. Thiagarajan, N. Caria, H. Schöder, et al., “Target volume delineation in oropharyngeal cancer: Impact of PET, MRI, and physical examination,” Int. J. Radiat. Oncol. Biol. Phys., vol. 83, no. 1, pp. 220–227, 2012.

[9] D. Bird, A. F. Scarsbrook, J. Sykes, et al., “Multimodality imaging with CT, MR and FDG-PET for radiotherapy target volume delineation in oropharyngeal squamous cell carcinoma,” BMC Cancer, vol. 15, no. 1, pp. 1–10, 2015.

[10] C. R. N. Rasch, R. J. H. M. Steenbakkers, I. Fitton, et al., “Decreased 3D observer variation with matched CT-MRI, for target delineation in Nasopharynx cancer,” Radiat. Oncol., vol. 5, no. 1, pp. 4–9, 2010.

[11] T. S. Hong, W. A. Tome, and P. M. Harari, “Heterogeneity in head and neck IMRT target design and clinical practice,” Radiother. Oncol., vol. 103, no. 1, pp. 92–98, 2012.

[12] V. Grégoire, P. Levendag, K. K. Ang, et al., “CT-based delineation of lymph node levels and related CTVs in the node-negative neck: DAHANCA, EORTC, GORTEC, NCIC, RTOG consensus guidelines,” Radiother. Oncol., vol. 69, no. 3, pp. 227–236, 2003.

[13] J. M. Michalski, C. Lawton, I. El Naqa, et al., “Development of RTOG Consensus Guidelines for the Definition of the Clinical Target Volume for Postoperative Conformal Radiation Therapy for Prostate Cancer,” Int. J. Radiat. Oncol., vol. 76, no. 2, pp. 361–368, 2011.

[14] A. W. Lee, W. T. Ng, J. J. Pan, et al., “International guideline for the delineation of the clinical target volumes (CTV) for nasopharyngeal carcinoma,” Radiother. Oncol., vol. 126, no. 1, pp. 25–36, 2017.

[15] V. Grégoire, K. Ang, W. Budach, et al., “Delineation of the neck node levels for head and neck tumors: A 2013 update. DAHANCA, EORTC, HKNPCSG, NCIC CTG, NCRI, RTOG, TROG consensus guidelines,” Radiother. Oncol., vol. 110, no. 1, pp. 172–181, 2014.

[16] V. Grégoire, M. Evans, Q. T. Le, et al., “Delineation of the primary tumour Clinical Target Volumes (CTV-P) in laryngeal, hypopharyngeal, oropharyngeal and oral cavity squamous cell carcinoma: AIRO, CACA, DAHANCA, EORTC, GEORCC, GORTEC, HKNPCSG, HNCIG, IAG-KHT, LPRHHT, NCIC CTG, NCRI, NRG Oncolog,” Radiother. Oncol., vol. 126, pp. 3–24, 2017.

[17] V. Grégoire, A. Eisbruch, M. Hamoir, et al., “Proposal for the delineation of the nodal CTV in the node-positive and the post-operative neck,” Radiother. Oncol., vol. 79, no. 1, pp. 15–20, 2006.

[18] S. R. de Mol van Otterloo, J. P. Christodouleas, E. L. A. Blezer, et al., “The MOMENTUM Study: An International Registry for the Evidence-Based Introduction of MR-Guided Adaptive Therapy,” Front. Oncol., vol. 10, Sep. 2020.

[19] H. M. Verkooijen, L. G. W. Kerkmeijer, C. D. Fuller, et al., “R-IDEAL: A Framework for Systematic Clinical Evaluation of Technical Innovations in Radiation Oncology,” Front. Oncol., vol. 7, Apr. 2017.

[20] H. Bahig, Y. Yuan, A. S. R. Mohamed, et al., “Magnetic Resonance-based Response Assessment and Dose Adaptation in Human Papilloma Virus Positive Tumors of the Oropharynx treated with Radiotherapy (MR-ADAPTOR): An R-IDEAL stage 2a-2b/Bayesian phase II trial,” Clin. Transl. Radiat. Oncol., vol. 13, pp. 19–23, Nov. 2018.

[21] B. A. McDonald, S. Vedam, J. Yang, et al., “Initial Feasibility and Clinical Implementation of Daily MR-Guided Adaptive Head and Neck Cancer Radiation Therapy on a 1.5T MR-Linac System: Prospective R-IDEAL 2a/2b Systematic Clinical Evaluation of Technical Innovation,” Int. J. Radiat. Oncol., Dec. 2020.

[22] L. R. Dice, “Measures of the Amount of Ecologic Association Between Species,” Ecology, vol. 26, no. 3, pp. 297–302, 1945.

[23] J. Yang, B. Haas, R. Fang, et al., “Atlas ranking and selection for automatic segmentation of the esophagus from CT scans,” Phys. Med. Biol., vol. 62, no. 23, pp. 9140–9158, 2017.

[24] S. K. Warfield, K. H. Zou, and W. M. Wells, “Simultaneous Truth and Performance Level Estimation (STAPLE): An Algorithm for the Validation of Image Segmentation,” vol. 23, no. 7, pp. 903–921, 2004.

[25] A. C. Riegel, A. M. Berson, S. Destian, et al., “Variability of gross tumor volume delineation in head-and-neck cancer using CT and PET/CT fusion,” Int. J. Radiat. Oncol., vol. 65, no. 3, pp. 726–732, Jul. 2006.

[26] C. M. Anderson, W. Sun, J. M. Buatti, et al., “Interobserver and intermodality variability in GTV delineation on simulation CT, FDG-PET, and MR Images of Head and Neck Cancer.,” Jacobs J. Radiat. Oncol., vol. 1, no. 1, p. 006, 2014.

[27] S. P. Ng, B. A. Dyer, J. Kalpathy-Cramer, et al., “A prospective in silico analysis of interdisciplinary and interobserver spatial variability in post-operative target delineation of high-risk oral cavity cancers: Does physician specialty matter?,” Clin. Transl. Radiat. Oncol., vol. 12, pp. 40–46, 2018.

[28] S. P. Ng, C. E. Cardenas, H. Elhalawani, et al., “Comparison of tumor delineation using dual energy computed tomography versus magnetic resonance imaging in head and neck cancer re-irradiation cases,” Phys. Imaging Radiat. Oncol., vol. 14, no. March, pp. 1–5, Apr. 2020.

[29] S. Gudi, S. Ghosh-Laskar, J. P. Agarwal, et al., “Interobserver Variability in the Delineation of Gross Tumour Volume and Specified Organs-at-risk During IMRT for Head and Neck Cancers and the Impact of FDG-PET/CT on Such Variability at the Primary Site,” J. Med. Imaging Radiat. Sci., vol. 48, no. 2, pp. 184–192, Jun. 2017.

[30] J. van der Veen, A. Gulyban, and S. Nuyts, “Interobserver variability in delineation of target volumes in head and neck cancer,” Radiother. Oncol., vol. 137, pp. 9–15, 2019.

[31] C. R. Hansen, J. Johansen, E. Samsøe, et al., “Consequences of introducing geometric GTV to CTV margin expansion in DAHANCA contouring guidelines for head and neck radiotherapy,” Radiother. Oncol., no. 126, pp. 43–47, 2018.

[32] C. E. Cardenas, R. E. McCarroll, L. E. Court, et al., “Deep Learning Algorithm for Auto-Delineation of High-Risk Oropharyngeal Clinical Target Volumes With Built-In Dice Similarity Coefficient Parameter Optimization Function,” Int. J. Radiat. Oncol. Biol. Phys., vol. 101, no. 2, pp. 468–478, 2018.

[33] C. E. Cardenas, B. M. Anderson, M. Aristophanous, et al., “Auto-delineation of oropharyngeal clinical target volumes using 3D convolutional neural networks,” Phys. Med. Biol., vol. 63, no. 21, 2018.

[34] J. Unkelbach, T. Bortfeld, C. E. Cardenas, et al., “The role of computational methods for automating and improving clinical target volume definition,” Radiother. Oncol., 2020.

[35] H. Ligtenberg, E. A. Jager, J. Caldas-Magalhaes, et al., “Modality-specific target definition for laryngeal and hypopharyngeal cancer on FDG-PET, CT and MRI,” Radiother. Oncol., vol. 123, no. 1, pp. 63–70, 2017.

[36] N. N. Chung, L. L. Ting, W. C. Hsu, et al., “Impact of magnetic resonance imaging versus CT on nasopharyngeal carcinoma: Primary tumor target delineation for radiotherapy,” Head Neck, vol. 26, no. 3, pp. 241–246, 2004.

[37] S. Adams, R. P. Baum, T. Stuckensen, et al., “Prospective comparison of 18F-FDG PET with conventional imaging modalities (CT, MRI, US) in lymph node staging of head and neck cancer,” Eur. J. Nucl. Med., vol. 25, no. 9, pp. 1255–1260, 1998.

[38] C. E. Cardenas, A. S. R. Mohamed, R. Tao, et al., “Prospective Qualitative and Quantitative Analysis of Real-Time Peer Review Quality Assurance Rounds Incorporating Direct Physical Examination for Head and Neck Cancer Radiation Therapy,” Int. J. Radiat. Oncol. Biol. Phys., vol. 98, no. 3, pp. 532–540, 2017.

[39] M. T. Ballo, G. M. Chronowski, P. J. Schlembach, et al., “Prospective peer review quality assurance for outpatient radiation therapy,” Pract. Radiat. Oncol., vol. 4, no. 5, pp. 279–284, 2014.

[40] E. A. Jager, H. Ligtenberg, J. Caldas-Magalhaes, et al., “Validated guidelines for tumor delineation on magnetic resonance imaging for laryngeal and hypopharyngeal cancer,” Acta Oncol. (Madr)., vol. 55, no. 11, pp. 1305–1312, Nov. 2016.

